# On the Use of Envelope Following Responses to Estimate Peripheral Level Compression in the Auditory System

**DOI:** 10.1101/2020.02.19.20024919

**Authors:** Gerard Encina-Llamas, Torsten Dau, Bastian Epp

## Abstract

Hearing impairment is a common and increasingly frequent problem among elderly people. The success of restoration or compensation therapies is strongly dependent on the development of reliable diagnostic methods for individual patients. The ability to compress the large level range of incoming sounds into a smaller range of vibration amplitudes on the basilar membrane (BM) is an important property of the healthy auditory system. Sensorineural hearing impairment typically leads to a decrease in sensitivity to sound and a reduction of the amount of compression observed in BM input-output functions in the cochlea. While sensitivity loss can be measured efficiently via audiometry, no measure has yet been provided that represents fast and reliable compression estimates in the individual listener. This would be useful to disentangle outer hair cells (OHC) from inner hair cells (IHC) damage. In the present study, magnitude-level functions obtained from envelope following response (EFR) to four simultaneously presented amplitude modulated tones were measured in normal hearing (NH) and sensorineural hearing impaired (HI) listeners. The slope of part of the EFR magnitude-level function was used to estimate level compression as a proxy of peripheral compression. The median values of the compression estimates in the group of NH listeners were found to be consistent with previously reported group-averaged compression estimates based on psychoacoustical measures and group-averaged distortion-product otoacoustic emission magnitude-level functions in human listeners. They were also similar to BM compression values measured invasively in non-human mammals. The EFR magnitude-level functions for the HI listeners were less compressive than those for the NH listeners. This is consistent with a reduction of BM compression. Given the numerical concordance between EFR-based compression estimates and group-averaged estimates from other methods, the frequency-specific (on-characteristic frequency (CF)) nature of BM compression was analysed through computer modelling. A computer model of the auditory nerve (AN) was used to simulate EFR magnitude-level functions at the level of the AN. The recorded EFRs were considered to represent neural activity originating mainly from the auditory brainstem-midbrain rather than a direct measure of AN activity. Nonetheless, the AN model simulations could account for the recorded data. The model simulations revealed that the growth of the EFR magnitude-level function might be highly influenced by contributions from off-CF neural populations. This compromises the possibility to estimate on-CF (i.e., frequency-specific or “local”) level compression with EFRs. Furthermore, the model showed that, while the slope of the EFR magnitude-level function is sensitive to a loss of BM compression observed in HI listeners due to OHC dysfunction, it is also sensitive to IHC dysfunction. Overall, it is concluded that EFR magnitude-level functions may not represent frequency-specific level compression in the auditory system.

## Introduction

Hearing impairment is one of the most common chronic troublesome conditions for elderly people in the ageing Western societies [1], and can imply a significant functional limitation [2]. Although cell regeneration techniques are under development [3, 4], its success is dependent on reliable and precise diagnostic measures that can differentiate between types of peripheral cellular damage in individual patients. A reliable estimate of cochlear compression could be used to assess differential damage of inner hair cells (IHC) and outer hair cells (OHC) [5]. An important characteristic of the healthy mammalian auditory system is the compressive transformation of the large dynamic range of input sound pressure levels to a narrower range of levels that can be processed by the sensory cells. Part of this compressive transformation is a consequence of the processing by the OHC in the cochlea. Although there is still some controversy about the precise mechanism underlying OHC function [e.g., 6, 7, 8, 9, 10], it is broadly accepted that OHC electro-motility provides a level-dependent gain to the movement of the basilar membrane (BM) in the healthy cochlea. This leads to a high sensitivity to low-level sounds and a compressive input/output (I/O) function [11] at the characteristic place of the BM for tonal stimuli. In addition to increasing the sensitivity, OHC function has also been associated with high frequency selectivity and a normal loudness growth with sound pressure level (SPL) [12].

Invasive physiological recordings in living non-human mammals allow precise measures of place-specific BM velocity-level functions using pure-tone stimuli [e.g., 13, 14, 15, 16, 17]. For a pure tone, the envelope of the resulting travelling wave shows a maximum at one specific cochlear place. Magnitude-level functions measured at or near this place (“on-characteristic frequency (CF)”) show a compressive growth of BM velocity with increasing SPL, consistent with the idea of a level-dependent amplification. Basal and apical to this place (“off-CF”), the magnitude-level functions show a linear growth [Figs. 6 and 7 in 15]. The combination of nonlinear on-CF and linear off-CF magnitude-level functions leads to a level-dependent BM excitation pattern with sharp tuning at low levels and broader tuning at higher levels. In the case of OHC dysfunction, on-CF magnitude-level functions show reduced compression. This leads to a lower on-CF response amplitude and a less level-dependent tuning of the BM excitation pattern [18]. Direct measurements of BM velocity are not possible in-vivo in humans due to ethical considerations. Instead, behavioural measurements using forward-masking paradigms [e.g., 19, 20, 21, 22, 23, 24] as well as objective measurements based on distortion product oto-acoustic emission (DPOAE) amplitudes [e.g., 25, 26] have been considered to estimate BM compression in humans.

Envelope following response (EFR) represent another objective and non-invasive measure to investigate auditory function. EFRs, also referred to as auditory steady-state response (ASSR), are gross electroencephalographic (EEG) potentials elicited by populations of neurons that fire synchronously (phase-locked) to the envelope of an acoustic stimulus. The EFR amplitude varies depending on the modulation frequency, with a predominant peak at 40 Hz (the so-called 40 Hz potential) and a smaller peak around 80-100 Hz (the 80 Hz potential; [27]). It has been suggested that, when the traditional clinical vertical electrode montage is used, EFRs to 80-100 Hz modulations are mainly generated by brainstem-midbrain sources, although cortical sources may also contribute [28, 29, 30]. EFRs to 40 Hz modulations are thought to have more dominant sources from cortical stages [31]. Due to their narrow bandwidth, multiple sinusoidally amplitude modulated (SAM) tones have been used to record EFRs evoked at multiple cochlear regions simultaneously to speed up clinical assessment [32, 33, 34]. While the carrier frequency of each SAM tone determines the cochlear region to be excited, different modulation frequencies produce different peaks in the recorded EEG spectrum [35], making it possible to separate the responses in the frequency domain. It has been demonstrated that EFRs can be recorded using four simultaneous SAM tones modulated between 80-100 Hz [33], a technique currently implemented in clinical systems to estimate hearing thresholds [e.g., 36]. At supra-threshold levels, EFR magnitude as a function of stimulus level (EFR magnitude-level functions) elicited from single SAM tones show a monotonic growth [37, 38]. It was shown that EFR magnitudes evoked by multiple SAM tones presented simultaneously, separated one octave apart and modulated around 80 Hz, grow monotonically up to about 60 dB SPL followed by a plateau [39]. EFR magnitudes are also influenced by the modulation depth. For a fixed stimulus level and modulation frequency, the magnitude of the EFR drops when reducing the modulation depth of the stimulus [37, 40]. Given that the modulation depth of a SAM tone is reduced when passed through a compressive nonlinearity (e.g., BM processing), the EFR magnitudes may be lower for a system with high compression compared to a system with low compression [see 41]. Consequently, the slope of the EFR magnitude-level function as a function of stimulus level should be shallower for a more compressive system compared to a less compressive system, potentially revealing the compressive growth of the BM.

In the present study, EFR magnitude-level functions in the frequency range of 80 Hz SAM tones were measured in normal hearing (NH) and hearing impaired (HI) listeners. Even though EFRs evoked by a 80 Hz modulation may be predominantly elicited at brainstem-midbrain auditory stages [42, 43, 31], the compression estimates derived from the slope of the EFR magnitude-level functions may reflect a combination of various compressive processes from the cochlea to the brainstem-midbrain, and even potentially at cortical levels [28, 29, 30]. Assuming that all sources of compression beyond cochlear processing are not or only minimally affected by OHC dysfunction, a change in BM compression will be reflected at more central levels in the auditory pathway (i.e., brainstem-midbrain). Such a change will then be represented in the magnitude of the EFR. The aim of the present study was thus to investigate whether peripheral level compression can be estimated simultaneously at four different carrier frequencies using EFR magnitude-level functions. It was anticipated that the amount of compression estimated through the EFRs will be higher in NH listeners compared to HI listeners due to the reduced contribution of BM compression in the HI listeners with sensorineural hearing loss. Prior to proposing the potential clinical use of the slope of the EFR magnitude-level functions as an estimate of peripheral compression, the stability of the measure must be demonstrated. EFR test-retest repeatability analysis was performed in NH listeners both to individual EFR data points and to EFR slopes. In addition to the analysis on the experimental data, a well-established phenomenological computer model of the auditory nerve (AN) [44] was used to investigate the mechanisms underlying the peripheral contributions to the EFRs generation. Due to the frequency-specific (on-CF) nature of BM compression, special attention was dedicated to analyse the effect of off-CF AN contributions to the compression estimates derived from the simulated EFR magnitude-level functions. Effects of OHC and IHC dysfunction on the EFR-based compression estimates were systematically investigated within the modelling framework.

## Methods

### Listeners

Twenty adult listeners (10 females, 34.0 ± 15.9 years) participated in this study. Listeners were separated into groups of 13 NH (8 females, 24 ± 3.2 years) and 7 HI (2 females, 56.2 ± 12.7 years). All NH listeners had audiometric thresholds below 15 dB hearing level (HL) at octave frequencies between 125 and 8000 Hz. All HI listeners were selected to have normal-hearing thresholds (≤ 20 dB HL) below 4000 Hz and a mild hearing impairment at 4000 Hz and above, with audiometric thresholds between 20 and 45 dB HL.

All participants provided informed consent and all experiments were approved by the Science-Ethics Committee for the Capital Region of Denmark (reference H-3-2013-004). The experiments were carried out in accordance with the corresponding guidelines and regulations on the use of human subjects. The listeners were economically compensated for participating in the experiments.

### Apparatus

The EFR recordings were performed in a dark, soundproof and electrically shielded booth, where the participants were seated in a comfortable reclined armchair. The participants were instructed to close their eyes and relax to avoid moving and were allowed to sleep. The recording and data analysis routines were implemented in MATLAB (The MathWorks, Inc., Natick, Massachusetts, USA). All acoustic stimuli were generated in MATLAB and presented using PLAYREC 2.1 (Humphrey, R., www.playrec.co.uk, 2008-2014) via a RME Fireface UCX soundcard (sampling rate *f*_s|sound_ = 48 kHz, 24 bits). The analogue acoustic signal was passed to a headphone buffer (HB7, Tucker-Davis Technologies) with a gain of −6 dB (stimulus levels > 55 dB SPL) or −27 dB (stimulus levels ≤ 55 dB SPL). The attenuated signal was presented through a pair of ER-2 insert earphones (Etymotic Research Inc.) mounted on an ER-10B+ low-noise DPOAE microphone probe (Etymotic Research Inc.) with ER10-14 foam eartips.

EFRs were recorded using a Biosemi ActiveTwo system (sampling rate *f*_s_|_EFR_ = 8192 Hz, speed mode 6, 24 bits). Five active pin-type electrodes were used. Three electrodes were mounted at positions P10, P9 (right and left extremes of the parietal coronal line) and Cz (vertex) following the 10-20 system [45]. The remaining two electrodes (common mode sense (CMS) and driven right leg (DRL)) were placed at the centre of the parieto-occipital coronal line (on either side of electrode POz). The electrodes CMS and DRL form a feedback loop that replace the “ground” electrode (the zero) in conventional EEG systems [46]. Conductive electrode gel was applied and the offset voltage was stabilised at < 20 mV for each electrode. The recorded EEG signals were hardware low-pass filtered by the EEG amplifier with a bandwidth limit of ⅕^th^ of the sampling frequency (anti-aliasing filter) and down sampled by a factor of 2 by the Biosemi software. The EEG data were stored to hard disk. The results shown in this study represent the Cz-P10 potential in response to right-ear stimulation, and the Cz-P9 potential in response to left-ear stimulation.

### EFR recordings

The EFR data were recorded in two sessions. In the first session (approx. two hours in duration), the EFR magnitude-level functions were recorded in the NH listeners using input levels in the range from 20 to 80 dB SPL, in steps of 5 dB. The second recording session (approx. 45 minutes in duration) took place on a different day usually about one month later than the first session. Three input levels (35, 55 and 70 dB SPL) were recorded again in the same NH listeners to evaluate the repeatability of the results. In all NH listeners, the right ear was stimulated. In the HI group, the multi-frequency recording was carried out in the level range from 30 to 80 dB SPL, in steps of 5 dB. Here, the recording ear was chosen depending on the individual listener’s audiogram, such that the amount of sensitivity loss due to the hearing impairment was as similar as possible within the group. There was no second recording session to evaluate repeatability for this subject group.

A multi-frequency stimulus consisting of four SAM tones was used. The SAM tones had carrier frequencies of 498, 1000, 2005 and 4011 Hz (referred to as 500, 1000, 2000 and 4000 Hz throughout this work) modulated at 81, 87, 93 and 98 Hz, respectively. The modulation depth was set to m = 85%. The four SAM tones were calibrated individually to the desired root mean squared (RMS) value using a B&K 4157 ear simulator, and were added later together (resulting in a final stimulus level that was 6 dB higher than that of each individual SAM tone). The stimuli were digitally generated as 1-s long epochs and continuously presented to the listener, where a trigger signal marked the beginning of a new epoch for later averaging. The total stimulus duration depended on the stimulus intensity to achieve a statistically significant EFR signal-to-noise ratio (SNR), based on a pilot study. Table 1 shows the stimulus duration used for each input level in the EFR recordings.

**Table 1.**
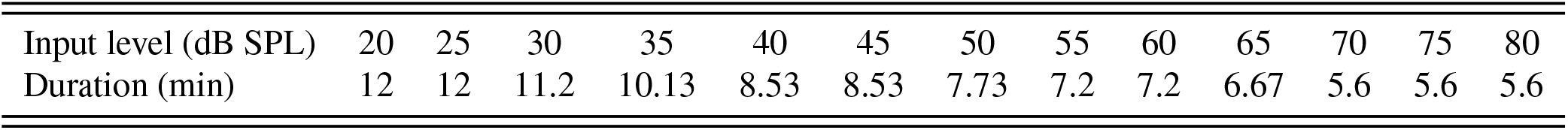
Duration of EFR stimuli for each input level used in the NH listeners.

The recorded EEG data were filtered using a fourth-order Butterworth digital band-pass filter with corner frequencies of 60 and 400 Hz, applied serially in forward and backward direction to yield zero phase. All recorded epochs with a maximum absolute amplitude that exceeded a voltage threshold of 80 µV in any of the channels were rejected to remove artefacts and noisy events from the average pool. Sixteen 1-s long epochs of EEG data were concatenated forming a trial to achieve a higher frequency resolution in the EFR spectrum analysis. In order to increase the SNR, the 16-s long trials were ensemble weighted averaged, where the inverse of the variance on each 1-s long epochs was used as weights [47].

A F-test was used to identify statistically significant responses by comparing the spectral power at the modulation frequency (EFR frequency) to the noise power in the range of 3 Hz below and above the modulation frequency [48, 27]. The power ratio (*F-ratio*) was calculated as the power in the EFR frequency bin divided by the averaged power in 3 Hz below and above the modulation frequency (96 bins). The probability (*p*) of the EFR power being different from the noise power can be calculated as 1 − *F*, with F representing the cumulative distribution function of the power ratio. The F-test was defined to be positive if *p* ≤ 0.01 (F critical value ≤ 4.8333, SNR > 5.84 dB), implying that the EFR frequency was statistically significant from the noise estimate. The F-test was custom implemented in MATLAB.

Compression was estimated from the slope of the EFR magnitude-level function. To estimate such slope, a piecewise linear function with two segments was used. The model was fitted to each individual EFR magnitude-level data using a non-linear least squares fitting method described by:

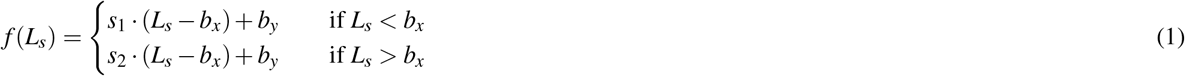

where *L*_*s*_ represents the stimulus input level, *s*_1_ the lower slope, *s*_2_ the upper slope and *b*_*x*_ and *b*_*y*_ represent the value on the abscissa and the ordinate at the break-point respectively. Motivated by the BM I/O function characteristics observed either in direct animal physiological recordings [e.g., 15] or used for human psychoacoustical estimates of cochlear compression [e.g., 19, 20, 12, 21], the lower slope was forced to be larger than the upper slope (i.e. *s*_1_ > *s*_2_), otherwise a single slope (first-order polynomial) was used. Moreover, if a single-slope model was found to provide a better fit than the two-slopes piecewise functions (based on an adjusted *R*^2^ statistic) for a given EFR magnitude-level function, this simpler model was used. Also, the two-slopes piecewise function was only considered if at least 3 significant data points were present on each segment of the fitting function. Only EFR readings significantly above the background noise floor were included in the fitting procedure. In order to test whether the estimated EFR slopes at two different frequencies were statistically different from each other, a two-sample permutation test for equality of the means was used [49, 50]. The test evaluates the hypothesis that the estimated EFR slopes for two given frequencies were a random partition of both frequency data added together, against the alternative hypothesis that the EFR slopes from one frequency were part of a population with a different mean than the other frequency. The test was performed using 100000 permutations using the Permute package implemented in Python [51].

The accuracy of the fitted slope to the EFR magnitude-level function will depend on the test-retest variability of each individual EFR data point. In order to have an estimate of the measurement variability, the repeatability of individual EFR magnitudes at three stimulus levels was assessed using a Bland-Altman analysis [52]. Data was also analysed using a one-way, random-effects, single-rater intra-class correlation (ICC) coefficient [53], as in [54], interpreted using the guidelines provided by [55]. Normality was ensured by means of a visual inspection of the corresponding quantile-quantile (Q-Q) plots [56] and by computing a Shapiro-Wilk normality test [57] (not shown). In the Bland-Altman analysis, the test-retest difference values were plotted against the mean response amplitude between two test runs. This method defines the limits of agreement (LoA) as 1.96 times the standard deviation of the differences. The method proposed by [58] was used to compute 95 % confidence interval (CI) for the mean of the differences and for the upper and lower LoA. The same repeatability analysis was performed on the EFR slopes between 35 and 55 dB SPL (Slope_55−35 dB_). The repeated EFR at 75 dB was not used for estimating the slope because it does not belong to the compressive part in the EFR magnitude-level functions. Slope_55−35 dB_ was estimated by computing the difference between the EFR magnitudes at each stimulus level divided by 20. Only statistically significant EFR data points (positive F-test) were considered in the repeatability analysis.

### AN model

A humanised phenomenological AN model [44] was used to simulate the activity of the AN. In short, the input acoustic stimulus waveform is processed by a linear filter mimicking the middle-ear filtering. The BM is modelled as a time-varying level-dependent filter-bank where a gain parameter models the effect induced by OHC motion. The IHC transmembrane potential is modelled by a rectifying non-linearity coupled to a low-pass filter that limits phase-locking. Fast and slow power-law functions are used to model offset adaptation and long-term adaptation observed in single AN unit recordings [59]. Short-term onset adaptation is modelled as an adaptive redocking mechanism with four synaptic release sites. The implementation of the AN model is similar to one used in [60]. Each simulation computes a total of 32000 AN fibres, distributed non-uniformly (with more density of fibres at mid CFs based on [61]) through 300 CFs (cochlear segments or IHCs) ranging from 125 Hz to 20 kHz. For each CF, a 61 % of high-spontaneous rate (SR) (HSR) fibres, 23 % of medium-SR (MSR) fibres and 16 % of low-SR (LSR) fibres were considered [62]. Hair-cell impairment was implemented by fitting the listener’s audiogram using the *fitaudiogram2* MATLAB function implemented by [59]. This function allows to define the proportion of threshold elevation that is attributed to either OHC or IHC dysfunction.

To simulate EFR magnitude-level functions at the level of the AN, throughout the manuscript referred to as EFR_AN_, the same stimulus as the one used in the recordings consisting of four simultaneous SAM tones was presented to the model but of a duration of 1.2-s. The stimuli were calibrated and presented to the AN model ranging from 5 to 100 dB SPL in steps of 5 dB. The spike trains obtained from the independently computed AN neurons for a given CF and fibre type were added together to obtain the summed AN activity at that CF, which is comparable to the peri-stimulus time histogram (PSTH) used to describe data from single neurons in experimental recordings. In order to analyse the steady-state encoding of a modulation, a 1-s long steady-state response, excluding onsets and offsets, was considered. A fast Fourier transform (FFT) was performed on the resulting synaptic output and the magnitude value at the modulation frequency bin was considered the simulated EFR_AN_.

In order to easily visualise which CFs may contribute to the total EFR_AN_ response, heatmap plots showing EFR_AN_ magnitude as a function of CF and stimulus levels were used (see Fig. 5a-g). In those plots, for each combination of CF and stimulus level, the colour represents the magnitude of the EFR_AN_ obtained from the frequency bin corresponding to the modulation frequency of interest (different for each carrier frequency) in the spectrum of the summed AN activity at that CF (the PSTH for that particular CF). The simulated EFR_AN_ magnitude-level functions (see Fig. 5h-k) are obtained by summing all the AN simulated activity across CFs and reading the magnitude at the modulation frequency bin from the spectrum of the summed PSTHs. For the analysis done at the on- and off-CF bands, the same procedure is performed over the summed AN activity of all CFs within the definition of on-CF band, or over the summed AN activity of all CFs except the ones of the on-CF band (off-CFs). The on-CF band was defined as the CFs ranging from 1/2-octave lower and ⅓-octave higher than the carrier frequency of the SAM tone (a fractional bandwidth of ≈ 28%), based on velocity-intensity functions recorded directly in the BM of non-human animal [15].

## Results

The data reported in this article are publicly available in a Zenodo repository [63].

### EFR magnitude-level functions

#### Normal-hearing listeners

Figure 1 shows the recorded EFR magnitude-level functions for one representative NH listener (NH01) for 500 Hz, 1000 Hz, 2000 Hz and 4000 Hz (panels a-d), respectively. The complete set of EFR data for all NH listeners is shown in Supplementary Fig. 1. All EFR magnitude-level functions were found to grow monotonically and compressively (with slopes <1 dB*/*dB) for stimulus levels between 20 and 50-65 dB SPL. The EFR magnitude-level functions obtained for the carrier frequencies 500, 1000 and 2000 Hz (panels a-c in Supplementary Fig. 1) showed a different trend than that obtained for the 4 kHz carrier (panel d). At 500, 1000 and 2000 Hz, the EFR magnitudes saturated, or slightly decreased, for stimulus levels above 50-65 dB SPL, leading to a break-point in the magnitude-level function. Figure 2b (blue symbols) shows box-plots indicating the fitted break-point levels at the four carrier frequencies. The median values for the break-point levels varied between 50 to 65 dB SPL. In contrast, no break-point was observed at 4 kHz (hence the absence of a NH box-plot at 4 kHz). At this frequency, the magnitude-level function was found to grow monotonically with a single slope (see also Supplementary Table 1). Figure 2a (blue symbols) shows the median values of the EFR slopes, which amounted to 0.24 dB/dB at 500 Hz, 0.31 dB/dB at 1000 Hz, 0.25 dB/dB at 2000 Hz and 0.21 dB/dB at 4000 Hz. The estimated EFR slopes in the NH listeners were not statistically different across frequency, except for the conditions at 1 kHz vs 4 kHz (Test statistic = 0.1418, *p* = 0.0277).

**Figure 1.**
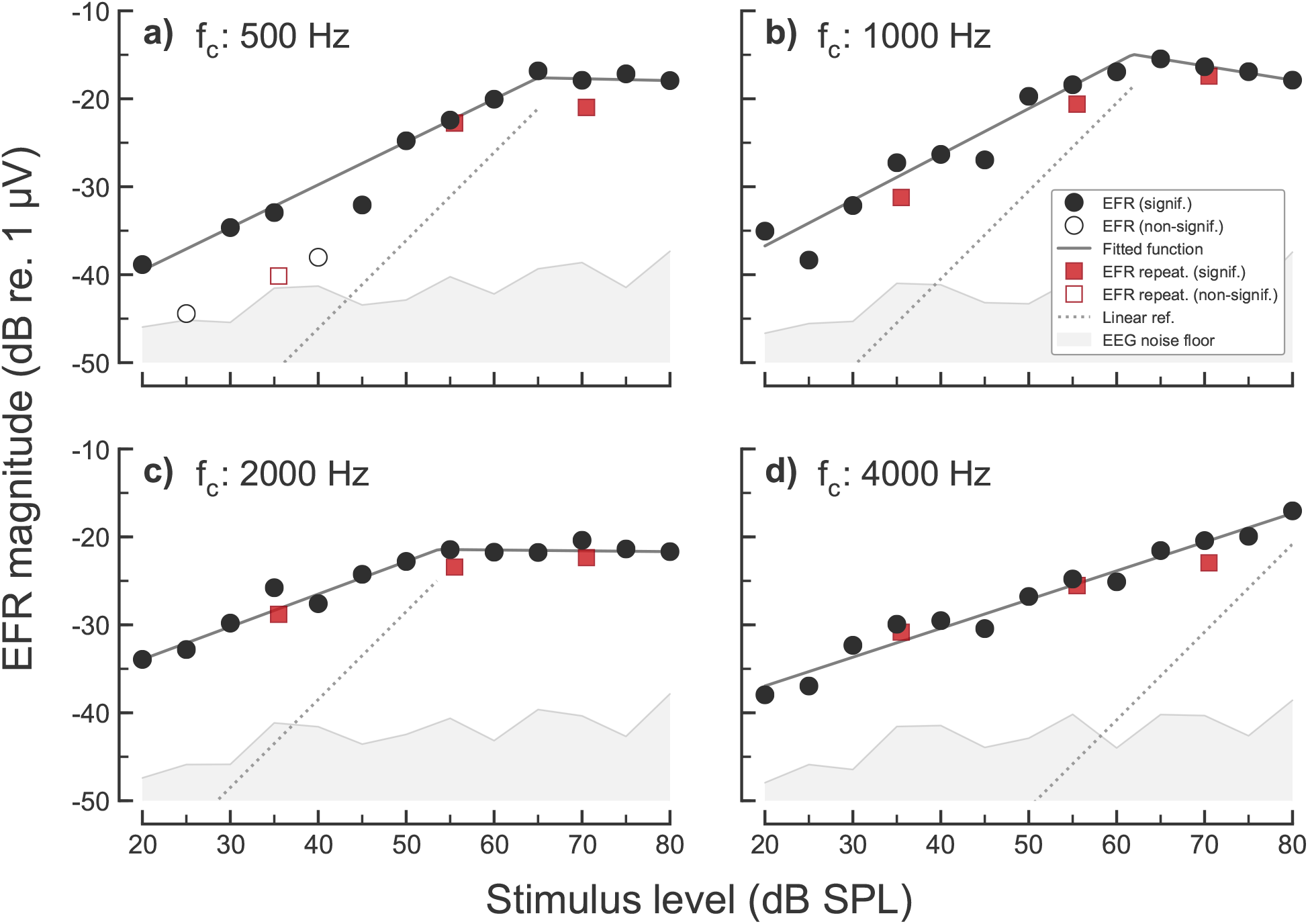
EFR magnitude-level functions recorded in one representative NH listener (NH01) for the carrier frequencies of a) 500 Hz, b) 1000 Hz, c) 2000 Hz and d) 4000 Hz. EFR magnitudes are represented as filled symbols in case of a statistically significant response (positive F-test) different from the EEG noise floor (grey shaded area). Open symbols show statistically non-significant (negative F-test) EFR magnitudes. Circles indicate the EFR magnitudes recorded in the first recording session of the test-retest repeatability, and red squares indicate EFR magnitudes recorded in the second recording session. Fitted curves to significant EFR data points are represented by the solid dark-grey functions. A linear reference with a slope of 1 dB*/*dB is indicated by the dotted line.

**Figure 2.**
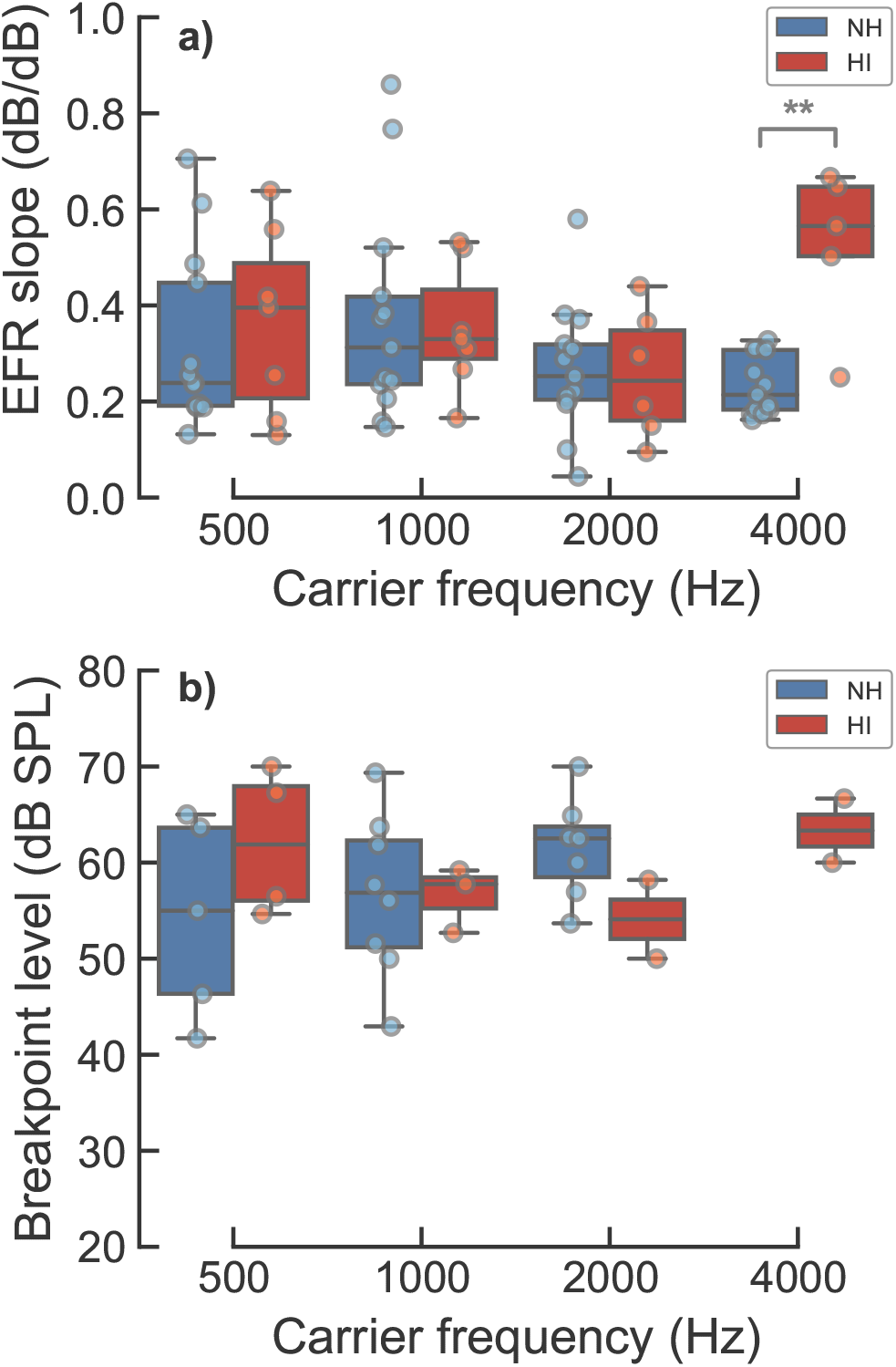
Fitted parameters to the EFR magnitude-level functions for the NH and HI listeners. Either a piecewise linear function with two segments or a single-order polynomial was fitted to the EFR data, selected based on adjusted *R*^2^ statistics. Panel a) shows box-plots with the fitted slopes obtained for the different carrier frequencies in the NH (blue) and HI (red) listeners. Panel b) shows box-plots with the fitted break-point levels for the different carrier frequencies when the two-slopes piecewise fit was used. The bottom and the top of each box represent the first and third quartiles, respectively, and the horizontal line inside each box represents the second quartile (the median). Whiskers indicate 1.5 times the interquartile range (IQR) of the lower and upper quartiles. The circles depict the raw observations. Statistical significance, based on a two-sample permutation test for equality of the means, is represented by the asterisks, where ** corresponds to *p* ≤ 0.01.

#### Hearing-impaired listeners

Figure 3 shows the EFR magnitude-level functions for one representative HI listener (HI01). The complete set of EFR data for all HI listeners is shown in Supplementary Fig. 2. The EFR magnitude-level functions for 500, 1000 and 2000 Hz carrier frequencies (panels a-d) showed similar trends as the ones observed for the NH listeners (shown in Fig. 1). At 4 kHz (Fig. 3d), the EFR magnitudes for stimulus levels up to 60 dB SPL were not statistically different from the EEG noise floor, whereas significant EFR magnitudes were obtained above 60 dB SPL for this particular listener, showing a compressive growth with level (slope < 1 dB/dB). This frequency is within the region of reduced sensitivity in this listeners’ audiogram (red arrow in panel d). Overall, the EFR magnitudes recorded in some of the HI listeners showed a lower SNR than in the NH listeners, resulting in a larger number of statistically non-significant data points (see Supplementary Table 2).

**Figure 3.**
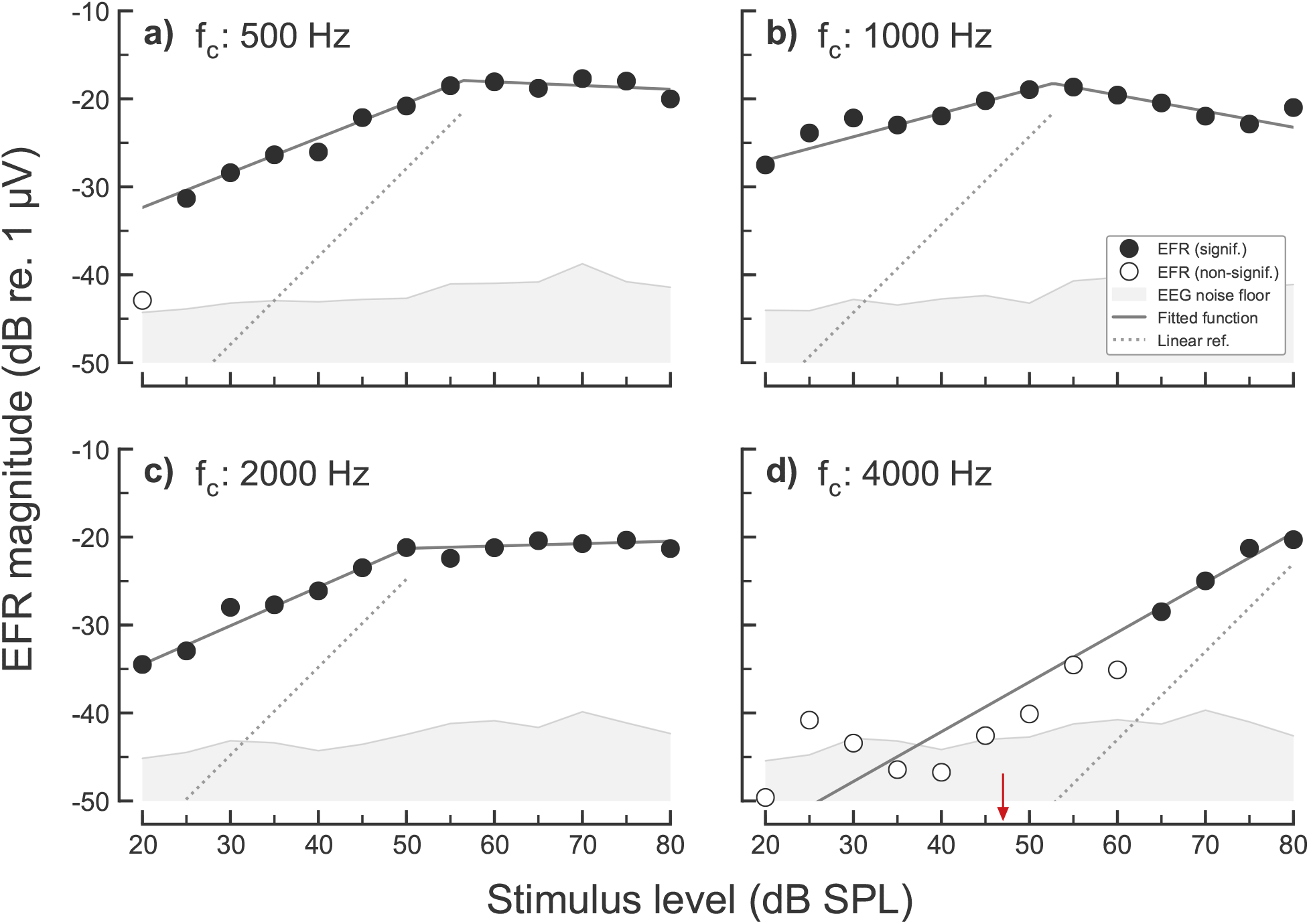
EFR magnitude-level functions recorded in one representative HI listener (HI01) for carrier frequencies of a) 500 Hz, b) 1000 Hz, c) 2000 Hz and d) 4000 Hz. Same representation as in Fig. 1 but not including the repeatability measurements. The small red arrow in panel d) indicates the behavioural hearing threshold of the listener at 4000 Hz in dB SPL.

The slopes of the EFR magnitude-level functions at 500, 1000 and 2000 Hz (i.e. the frequencies where all listeners were considered to have “normal” audiometric thresholds) were not statistically different between the NH and the HI listeners (see Fig. 2a). In contrast, the EFR slopes at 4 kHz were significantly steeper (higher values) for the HI listeners than for the NH listeners (Test statistic = 0.2933, *p* = 0.0012). The median values of the EFR slopes for the HI listeners were 0.40 dB/dB, 0.33 dB/dB, 0.24 dB/dB and 0.57 dB/dB for the carrier frequencies 500, 1000, 2000 and 4000 Hz, respectively.

### EFR test-retest repeatability

Figure 4 shows the main results of the repeatability analysis of individual EFR magnitudes and the EFR slopes, following the Bland-Altman method [52]. Panels a-b) show the repeatability analysis on the EFR slopes between stimulus levels of 35 and 55 dB SPL (Slope_55−35 dB_). Panels c-d) show the repeatability analysis on individual EFR magnitudes pooled across all frequencies and stimulus levels. Panels e-h) show the repeatability analysis of individual EFR magnitudes for each carrier frequency (from 500 Hz to 4 kHz, respectively). Panel a) shows the relation between Slope_55−35 dB_ in the test and retest conditions. The ICC coefficient was 0.219 (−0.34, 0.67), indicating a “poor” correlation. Panel b) shows a Bland-Altman plot of the same data. The solid black horizontal line indicate the mean of the differences and the grey band surrounding it indicate its 95 % CI. The mean of the slope difference was 0.03 dB*/*dB, indicating no systematic bias. The dashed lines show the upper and lower LoA, amounting 0.34 dB*/*dB and −0.27 dB*/*dB, respectively, with its 95 % CI depicted as grey surrounding areas.

**Figure 4.**
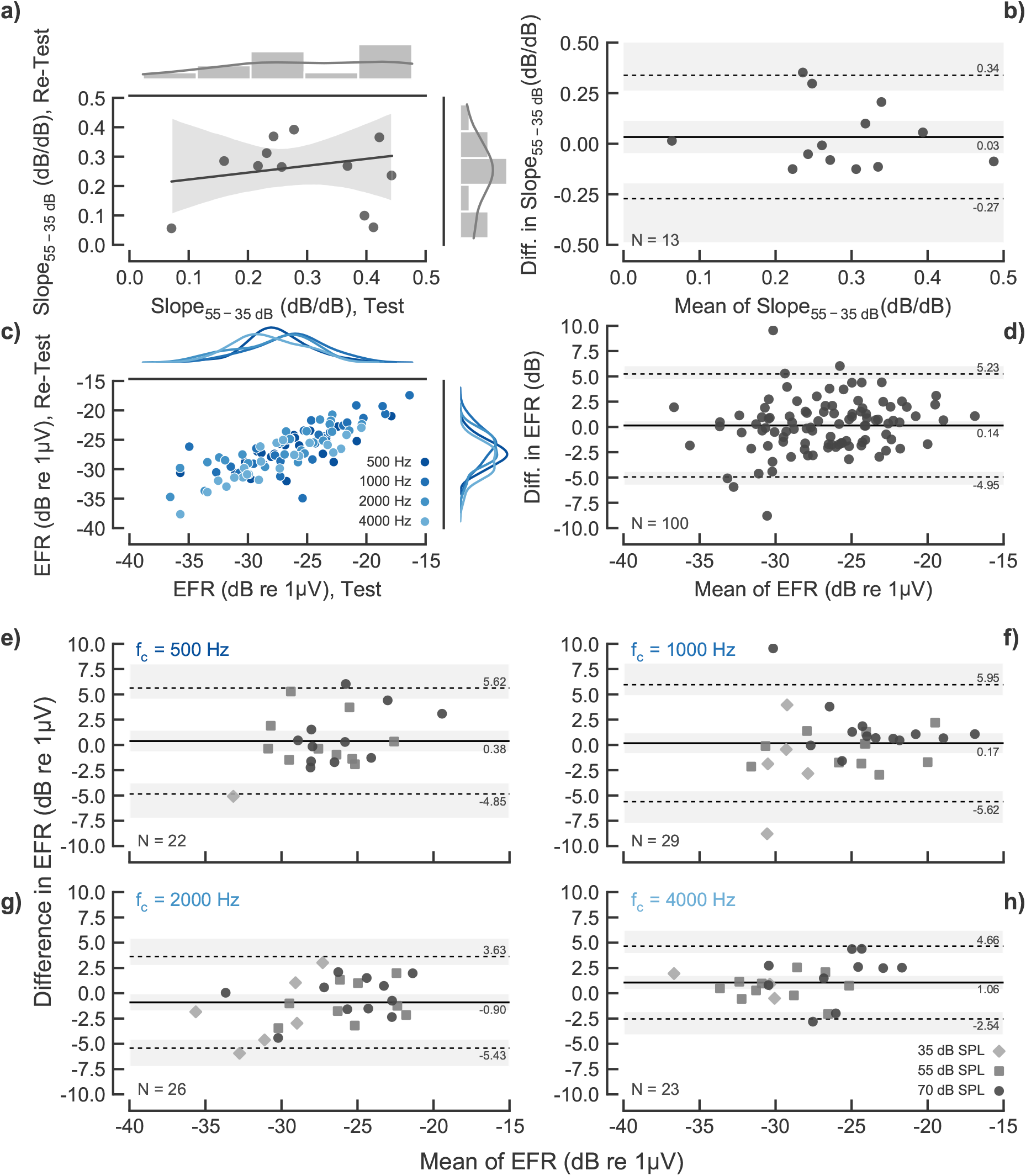
Test-Retest repeatability analysis of the EFR magnitudes and the EFR slopes in the NH listeners. Panel a) show the relation between the test and retest data of the EFR slope. Top and right margins show a histogram and kernel density function of the data. The black-solid line and grey area in the main plot show the fit of a linear regression model and its 95 % CI. Panel b) shows the difference of the test-retest EFR slopes as a function of the test-retest mean (a Bland-Altman plot). The solid-black horizontal line show the mean of the differences. The dashed-grey horizontal lines show the upper and lower LoA. The corresponding values are indicated in the right side of each line, while the grey semitransparent bands show the 95 % CI for each line. Panel c) show the relation between the test and retest EFR magnitudes pooled across carrier frequency and level. EFR values for each carrier frequency are indicated as different intensities of blue colour. Top and right margins show kernel density functions of the distribution of the data. Panel d) shows a Bland-Altman plot of the EFR test-retest data pooled across frequencies and levels. Panel e-h) show Bland-Altman plots of the EFR test-retest data for the carrier frequencies of 500 Hz, 1 kHz, 2 kHz and 4 kHz, respectively. Different markers indicate the stimulation level of each EFR data point, as shown in the legend in panel h). Only statistically significant EFRs were considered in the repeatability analysis, indicated by the N value in the bottom-left part of each Bland-Altman plot.

This indicates also the poor repeatability of the EFR Slope_55−35 dB_, as repeating a measurement of the EFR slope can lead to variations as large as about ± 0.3 dB*/*dB. Panel c) shows the relation between individual EFR test and retest magnitudes agnostic to carrier frequency and stimulation level. Carrier frequencies are indicated as different intensities of blue colour. The top and right margins show the kernel density function of the distributions of each frequency, showing the absence of a frequency-dependent structure in the data. The ICC coefficient was 0.792 (0.71, 0.86), indicating a “good” correlation. Panel d) shows the corresponding Bland-Altman analysis. The mean of the difference in EFR magnitudes (0.14 dB) does not show a systematic bias, and the LoAs indicate that the EFR test-retest repeatability is of about ± 5 dB. The analysis performed segmenting the data by carrier frequency (panels e-h) show a similar EFR test-retest repeatability for all carrier frequencies, with a slightly lower LoA for the 4 kHz carrier. The ICC coefficients for each carrier frequency were: 0.679 (0.38, 0.85) at 500 Hz, 0.751 (0.54, 0.87) at 1 kHz, 0.816 (0.63, 0.91) at 2 kHz and 0.848 (0.68, 0.93) at 4 kHz; interpreted as “good” repeatability for all frequencies except at 500 Hz, which is “moderate”. The repeatability results obtained in this study were similar to those presented in previous studies [64, 65, 54], even though the EEG recording systems, stimuli and listeners differed across studies.

### AN model simulations

Considering that BM compression is a frequency-specific (on-CF) phenomenon [15], a model of the AN was used to investigate the peripheral contributions to the EFR from each CF. Figure 5 shows simulated neural activity derived from the AN model by [44] in response to the same four SAM tones (rows) as considered in the experimental recordings. Simulated envelope-based AN activity (for details regarding the calculation of EFR_AN_ from the AN model output, see the Methods section), both for NH and HI are presented as a function of CF and stimulus level, as well as a summed across CFs. The AN responses for the carrier frequencies 500, 1000 and 2000 Hz were similar for the NH and the HI simulations (panels a-f), resulting in almost identical EFR_AN_ magnitude-level functions (panels i-k). At 4 kHz, EFR_AN_ magnitudes for the HI simulations (panel h and red diamonds in panel l) were not statistically significant from the noise floor at low stimulus intensities, consistent with a threshold elevation. Here, at threshold (i.e., at about 30-40 dB SPL), AN neurons tuned to a broader range of CFs responded phase-locked to the modulation frequency compared to the very narrow range of CFs in the case of the NH simulations (panel g and blue function in panel l), that showed a threshold at about 0 dB SPL. This is consistent with the broadening of frequency tuning observed in AN neurons in cochlear regions with OHC dysfunction [66]. Thus, hair-cell dysfunction led to abnormal EFR_AN_ magnitude-level functions, with non-significant responses at low input levels, followed by a steeper (less compressive) growth function at medium input levels that converged towards the compressive growth observed in the NH case (blue circles) at higher input levels.

The broadening of the range of contributing AN neurons with increasing stimulus level was found for all carrier frequencies (panels a-h). For each carrier frequency, the AN activity is limited to a narrow on-CF region at low stimulus levels (indicated by the horizontal orange-dashed lines in panels a-h). With increasing stimulus level, the range of AN activity broadens towards off-CF regions due to the recruitment of neurons tuned to higher CFs. This broadening is continuous for the 4 kHz carrier whereas there is a saturation in the EFR_AN_ magnitude-level functions (panels i-k) obtained for the three lower-frequency carriers due to an interference between the neural activity of a higher frequency carrier onto a lower frequency carrier.

Solid blue and red lines in panels i-l) in Fig. 5 show the growth of the BM output in the model for NH and HI, respectively, for each carrier frequency. The BM magnitude-level functions show a linear growth at low stimulus levels that bends towards a compressive growth at medium-to-high levels. This is less clear at 500 Hz because the BM gain in the model is lower at this frequency [68]. With a mild hearing impairment (panel l) the BM output shows reduced sensitivity at low stimulus levels but residual compression at higher levels. The compression values estimated from the output of the BM in the model are relatively similar to the compression values estimated from simulated EFR_AN_ magnitude-level functions. The same piecewise linear function with two segments as used in the experimental EFR data was used here for the model simulations. The compression values from the simulated NH BM output amounted 0.62, 0.33, 0.26 and 0.27 dB*/*dB at the four carrier frequencies, respectively; versus 0.58, 0.34, 0.39 and 0.23 dB*/*dB at the same frequencies for the EFR_AN_ magnitude-level functions. In the case of simulated HI, BM compression values were 0.66, 0.33, 0.27 and 0.32 dB*/*dB versus 0.59, 0.32, 0.36 and 0.25 dB*/*dB for the EFRs_AN_. Despite the similarity, one should bear in mind that the BM output for a given carrier frequency reflects the response of the cochlear segment tuned to that specific frequency, i.e., an on-CF response; whereas EFR_AN_ magnitude-level functions result from the addition of any CF responding to the modulation frequency, as shown in colour gradient in panels a-h in Fig. 5. A comparison between on- and off-CF contributions to the EFR_AN_ can be found in the Discussion section.

## Discussion

### Compression estimates based on EFR magnitude-level functions

The slopes of the experimental EFR magnitude-level function for the NH listeners varied between 0.2 and 0.35 dB*/*dB, and were not statistically different from the slopes at the non-impaired frequencies (500, 1000 and 2000 Hz) for the HI listeners (see Fig. 2a). On a group average, these values are similar to BM compression estimates obtained using direct invasive methods in healthy non-human animal models, and with non-invasive physiological and behavioural compression estimates in NH humans. Compression values estimated from the slope of BM velocity-intensity I/O functions in chinchillas at medium-to-high stimulation levels (40-90 dB SPL) varied between 0.2 and 0.5 dB*/*dB [15]. Group-averaged psychoacoustical compression estimates in NH humans were found to be between 0.15 to 0.35 dB*/*dB [e.g., 19, 20, 69, 70, 71, 72, 73]. Compression estimates using group-averaged DPOAE magnitude-level functions were shown to be of about 0.2 dB*/*dB in NH listeners at moderate stimulus levels (50 to 70 dB SPL) [e.g., 26, 25, 74]. Regarding the EFR recordings obtained in the HI listeners in the current study, some of the characteristics in the data also seem consistent with the results obtained with the other methods used to estimate cochlear compression. The slopes at 4 kHz (where the HI listeners showed a mild hearing loss) were significantly higher than the corresponding slopes for the NH listeners. The steeper growth function in the HI listeners is consistent with the concept of reduced BM compression observed or estimated with other methods [e.g., 18, 19, 20, 74, 75]. In addition, the increase of the lowest stimulus level at which an EFR could be measured in the HI listeners is consistent with the corresponding increased pure-tone threshold at that frequency [see also, 37, 76, 77, 78, 79, 80, 81]. Thus, based on the group-averaged numerical values, the similarity of the compression estimates obtained across the different methods, including the EFR, may suggest that similar aspects of peripheral auditory processing are reflected in the different measures.

However, while the slope value of the compressive part of the respective level-growth functions is similar across methods, the overall shape of the magnitude-level functions differs. For example, while a change in the slope of the magnitude-level functions (often referred to as a “break-point” or “knee point”) can be identified both in the EFR results as well as in the behavioural measures, there are substantial differences. In fact, in the behavioural studies, a break-point has typically been estimated [e.g., 20, 69, 82, 12, 72] at stimulus levels at or below about 45 dB SPL, whereby the slope of the estimated BM I/O function has been usually fitted to be close to one at lower levels, reflecting a linear growth. The slope beyond the break-point, at medium-to-high levels, has commonly shown a compressive growth in NH listeners. In contrast, for the EFR magnitude-level functions obtained in the present study, no linear growth was found, in any of the listeners at any frequency, at the lowest levels considered. While this characteristic of the EFR magnitude-level seems inconsistent with the behaviourally estimated BM I/O functions, it may not be inconsistent with data from non-human animal recordings which show that a linearised BM growth with input level can occur at stimulus levels below 20 dB SPL [see Fig. 3 in 15], which were input levels not tested in the present EFR study. In addition, the simulated EFR_AN_ magnitude-level functions showed growths close to 1 dB*/*dB only at very low stimulus levels below 30 dB SPL (panels i-l in Fig. 5), impossible to see from the recorded data. Furthermore, the experimental EFR magnitude-level functions showed a break-point at about 50-65 dB SPL (see Fig 2b); i.e., at higher levels than in the behavioural studies. This break-point actually reflected the transition between the compressive growth at low-medium stimulus levels and the level region where the EFR magnitudes saturated (at 500, 1000 and 2000 Hz), consistent with a previous study [39]. Thus, the characteristic of the EFR magnitude-level function, including its compressive behaviour, do not seem to reflect the same processes that underlie the behaviourally estimated BM I/O function. The same may hold in relation to the level-growth functions obtained with non-invasive physiological methods such as DPOAEs as well as the invasive (non-human) measures, as further outlined below.

**Figure 5.**
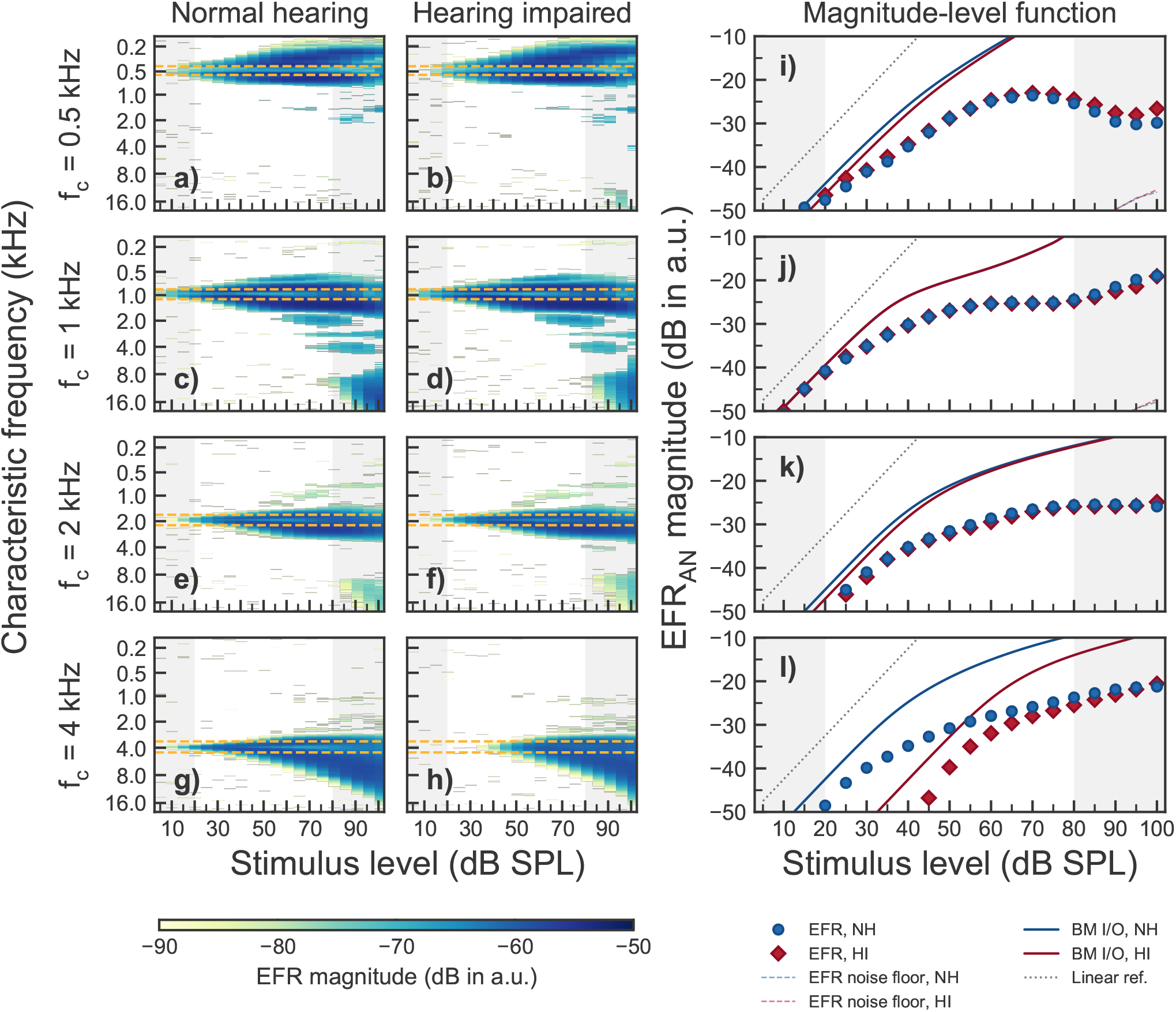
Simulated EFRs at the level of the AN (EFR_AN_) obtained with four simultaneously presented SAM tones as the ones used in the experiments. Panels a-h show EFR_AN_ magnitudes as a function of stimulus levels and CF for each of the four carrier frequencies (rows). The colour gradient indicate significant EFR_AN_ magnitudes. The horizontal orange dashed lines indicate the on-frequency range as defined in [15]. The vertical grey shaded areas show input level ranges outside the level ranges considered in the experiments. The two leftmost columns show simulated EFR_AN_ for the mean audiogram of the NH (left) and HI listeners (middle), respectively. Panels i-j in the rightmost column show the corresponding EFR_AN_magnitude-level functions obtained by summing up all AN activity across CF. Blue circles show NH simulations and red diamonds represent HI simulations. The thin blue and red dotted lines represent noise floor estimates (only visible in panels i and j). The blue and red solid lines show the BM output of the model for NH and HI, respectively. The black dotted lines show a linear reference. A combination of 2/3 of OHC dysfunction and ⅓ of IHC dysfunction [5, 67] was assumed to adjust the AN model parameters to account for the mean audiogram values in each listener’s group.

The repeatability analysis of individual EFR recordings showed a “good” test-retest repeatability based on the ICC method. The Bland-Altman method indicated that EFRs can fluctuate between about ± 5 dB from session to session. However, the test-retest repeatability of the EFR slope between 35 and 55 dB SPL in the present study was “poor” due to error propagation, as more than one EFR data points are used for estimating the slope. This is problematic for a potential clinical use of the EFR slope. The repeatability results of individual EFRs were consistent with other analyses from literature. Repeatability of individual EFR magnitudes were reported to be ICC = 0.93 (“excellent”) and ICC = 0.71 (“good”) for fully modulated (m = 100%) and shallow modulated (m = 50%) EFRs, respectively [54], leaving the repeatability coefficient in the present study in between (ICC = 0.792) for EFRs with modulation depths of m = 85%. Similarly, the Pearson correlation was reported to be *r* = 0.91 in another study that used single carriers modulated at m = 100% presented at 50 dB HL [65]. Moreover, the LoA of EFRs elicited also by a single carrier frequency modulated at m = 100% presented at 50 dB HL were found to be of about ± 40 % of the mean linear EFR amplitude, corresponding to about ± 3 dB [64], in comparison to the about ± 5 dB in the present study.

### On-versus off-CF contributions to compression estimates

The compressive growth of BM I/O functions measured locally in animal models reflects on-CF responses at a narrow BM range. At off-CF places, BM I/O functions have been demonstrated to grow linearly [see Figs. 6 and 7 in 15]. Thus, in order to estimate on-CF (i.e., place- or frequency-specific) compression using EFRs (or any other method), the response needs to be dominated by such on-CF processing. At low intensities, a narrow-band stimulus (e.g., aSAM tone) excites a narrow region of the BM and the AN. Thus, the EFR responses are likely to be dominated by the activity of a small population of neurons tuned to the centre frequency of the stimulus. However, at medium and high stimulus levels, the excitation pattern on the BM broadens and a larger population of AN neurons tuned to frequencies remote from the centre frequency contribute to the gross activity. Indeed, based on simulations of neural activity at the level of the AN using the model by [59, 83], it was shown that responses to a single SAM tone presented at medium-to-high stimulus levels are dominated by contributions from off-CF HSR fibres [60]. This is despite the fact that the maximum of the BM excitation is located at on-CF.

**Figure 6.**
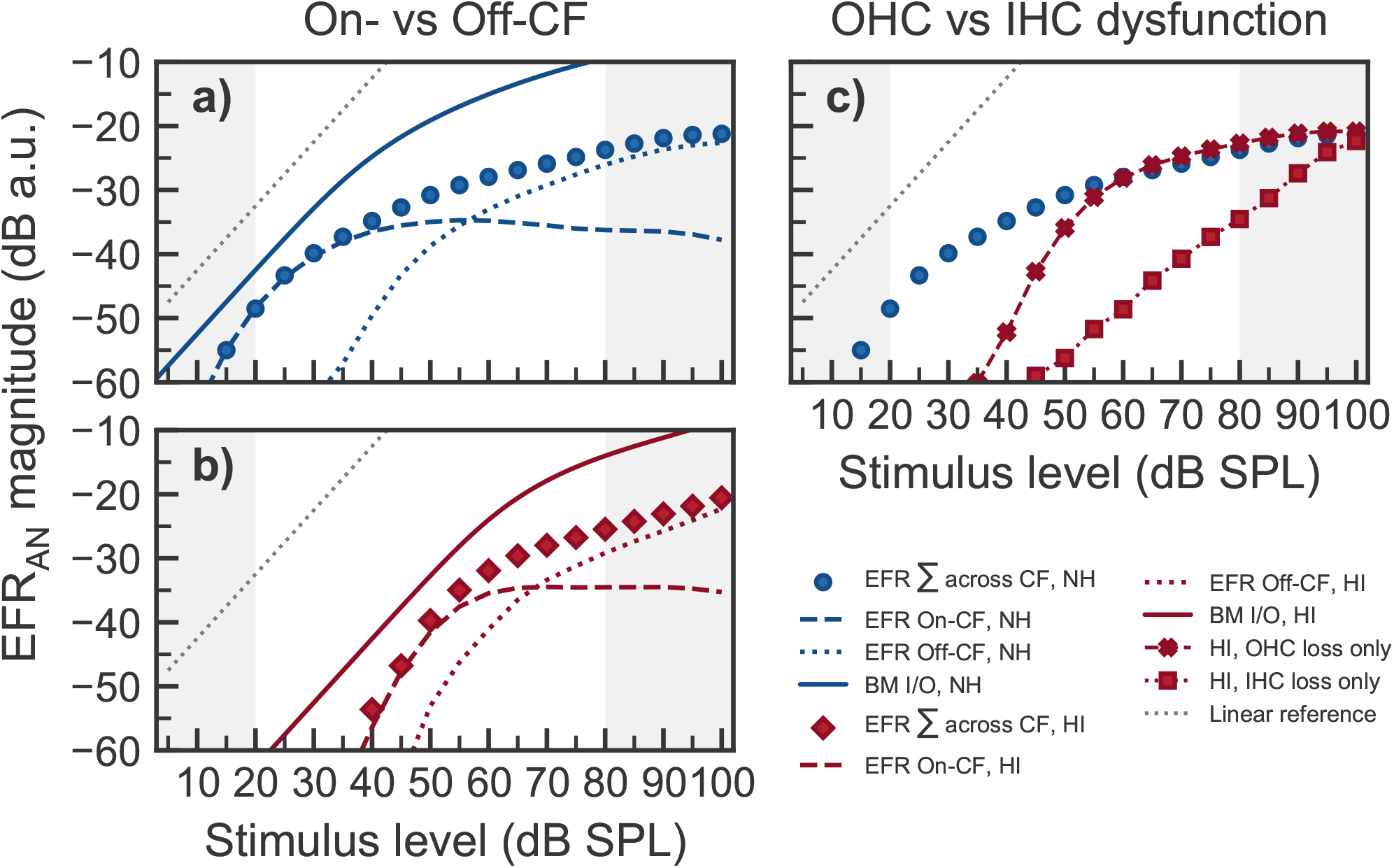
Analysis of on-vs off-CF contributions and OHC vs IHC dysfunction on the simulated EFR_AN_ magnitude-level functions from the 4 kHz component. Panel a) shows NH simulated EFR_AN_ magnitude-level function after summing contributions across CF (circles), the contribution to EFR_AN_ from neurons in the on-CF range (dashed line) and the contributions from off-CF neurons (dotted line). The solid line shows the growth of the BM output. Panel b) shows the same as panel a but assuming hearing impairment with a combinations of 2/3 of OHC dysfunction and ⅓ of IHC dysfunction. Panel c) shows the same simulated HI EFR_AN_ magnitude-level functions as in panel b) (summed across CFs) but assigning the threshold elevation to only OHC dysfunction (red crosses, dashed line) or to only IHC dysfunction (red squares, dotted line). The NH simulation is shown by blue circles as a reference. The grey-dotted line in all panels indicate a linear growth.

When presenting more then one SAM tone simultaneously, the presence of a SAM tone of higher carrier frequency prevented a SAM tone at a lower frequency to recruit AN neurons tuned to higher CFs (Fig 5, see panels a-f). This was not the case for the 4 kHz component (panels g and h), where off-CF basal neurons could be recruited without interference from another SAM tone. This is supported by findings from invasive recordings in non-human animals showing that AN fibres can follow the periodicity of a high level tone with frequency energy below the CF of the fibre [e.g., 84, 85]. Also, EFRs recorded in rats have shown the effect of a second, high-frequency SAM carrier onto the encoding of a low-frequency SAM carrier [86]. Consistent with the model simulations in the present study, [86] concluded that the presence of the second SAM tone basal to the place of the main low-frequency SAM tone caused a reduction of the EFR to the lower-frequency component due to reduced recruitment of AN fibres located basally relative to the on-CF location. Such interaction of a high-frequency SAM carrier onto the AN activity induced by a lower-frequency carrier is the reason underlying the saturation of the simulated EFR_AN_ magnitude-level functions above 50-60 dB dB SPL, as observed at 500, 1000 and 2000 Hz (panels i and j in Fig. 5). At 4 kHz, the model shows a monotonic growth of EFR_AN_ magnitudes at stimulus levels above 30 dB SPL in the NH simulations (panel l in Fig. 5). This is fully consistent with the single-slope growth function observed in the experimental EFR data (panel d in Fig. 1 and panel b in Fig. 2). This is because off-CF basal neurons could be recruited at high stimulus levels without interference, strongly contributing to the compound response. Similarly, simulated EFR_AN_ magnitude-level functions using a single SAM tone at 2 kHz resulted also in non-saturating (monotonically increasing) growth functions (see Fig. 4a in [60]). Thus, the model offers a comprehensive explanation for the saturation of the EFR magnitude-level functions observed at 500, 1000 and 2000 Hz in the recorded data (see panels a-c in Fig. 1 and Fig. 3).

The orange-dashed lines in Fig. 5 (panels a-h) indicate the limits of the on-CF range as derived from direct BM recordings in non-human mammals [15]. Within this on-CF range, the EFR_AN_ magnitude first increases with input level, shows a maximum at about 25-30 dB above threshold, and then decreases with increasing stimulus level. This is more clearly shown in the dashed-blue function in Fig. 6a. At medium-to-high stimulus levels, the modulation (and therefore the EFR_AN_) was found to be more dominant at off-CFs than at on-CFs (see also panels a and b in Fig. 6). At these higher levels, many on-CF HSR fibres will saturate in rate and do not strongly encode the modulations. In contrast, AN neurons tuned to off-CFs (including HSR fibres) are still excited below saturation level, due to its lower off-CF sensitivity (as reflected in their tuning curves). Thus, the off-CF HSR neurons more robustly encode the amplitude modulations at these higher levels. This is consistent with physiological recordings from the cat AN showing that off-CF AN fibres exhibit higher synchrony to high intensity SAM tones than on-CF AN fibres [87]. Nonetheless, the EFR_AN_ magnitude-level function obtained after summing across CF continued to grow monotonically due to a further increase of off-CF contributions. Panels a) and b) in Fig. 6 show the overall contribution of on-and off-CFs to the compound response to the 4 kHz SAM tone. The NH simulations are shown in blue and the HI simulations are indicated in red. The compressive slope estimated by fitting the piecewise function to the EFR_AN_ magnitude-level function (blue circles in panel a) results from the mixture of on- (dashed-blue line) and off-CF (dotted-blue line) contributions. The solid-blue line shows the BM output at 4 kHz, which reflects a purely on-CF response, and hence shows a compressive growth at medium-to-high stimulus level. At the same level range, the corresponding EFR_AN_ on-CF response (dashed-blue line) shows a decreasing function with increasing level, consistent with physiological results in non-human animals [87]. For this particular carrier frequency, the compression value from the simulated BM was 0.27 dB*/*dB, very similar to the 0.23 dB*/*dB compression value from the total simulated EFR_AN_ magnitude-level function. But it was completely different from the slightly negative growth of −0.02 dB*/*dB from the on-CF EFR_AN_, and similar to the 0.29 dB*/*dB growth from the off-CF EFR_AN_. The same effect can be seen in the HI simulations (see Fig. 6b). Therefore, while BM compression purely reflects on-CF processing, the estimates of compression obtained from EFR magnitude-level functions do not exclusively reflect on-CF cochlear compression, because its generation in the AN is strongly influenced by off-CF contributions, according to the model. This is consistent with the limitations of estimating place-specific cochlear dispersion using EFRs [88].

IHC dysfunction (but intact OHC function) is considered to lead to a BM I/O function with comparable compression estimates as in the NH listeners [e.g., 69]. Figure 6c shows simulations of the 4 kHz EFR_AN_ magnitude-level function when accounting for the mild threshold elevation with only OHC dysfunction (red crosses, dashed line) or with only IHC dysfunction (red squares, dotted line). Even though both types of hair-cell impairment led to a similar threshold elevation of 30-40 dB SPL, the growth function showed different shapes. In the case of only OHC dysfunction, EFRs_AN_ grew steeply just above threshold and became as compressive as in the NH case (blue circles) at levels beyond about 60 dB SPL. This steeply growing part, dominated by on-CF processing (see dashed-red line in panel b), resulted as a consequence of the loss of local on-CF gain due to simulated OHC dysfunction. The compressive growth that follows it at levels beyond 60 dB SPL resulted from the off-CF processing of the 4 kHz SAM tone by basally tuned AN neurons (i.e., off-CF contributions). These neurons are not affected by OHC dysfunction because they are excited through their tails [66]. In the case of only IHC dysfunction, processing of both on- and off-CF AN neurons was affected, as their whole tuning curve shifts towards higher levels [66]. Even though cochlear gain was not reduced (see the sharply tuned response in Supplementary Fig. 4h) and residual BM compression was “normal” (see the solid-red line in panel b), this resulted in an EFRs_AN_ function increasing monotonically with a mildly compressive single slope (red squares, dotted line in Fig. 6c). Therefore, the reduction of BM compression due to OHC dysfunction cannot be extracted from the simulated EFRs_AN_ magnitude-level functions because of the dominance of off-CF contributions at higher stimulus levels.

The assumption of reflecting frequency-specific local compression might also be challenged in the case of other measurement paradigms, such as those based on DPOAEs or psychoacoustical masking paradigms. The slope of DPOAE magnitudes-level functions was proposed to be used as an estimate of local BM compression [25]. The distortion source of the emission is usually simplified as a single source located at the peak of the travelling wave envelope (at on-CF), although many distortion sources might be induced in the region where the travelling waves of the two primaries, the pair of pure tones that induce the emission, overlap [89]. This overlap strongly depends on the level and frequency ratio of the primaries. The extent of potential off-CF contributions to the non-linear component of the DPOAE at high stimulus levels is not yet fully understood [90], which could be a potential confound in the estimate of BM compression. Regarding behaviourally obtained estimates of BM I/O functions, on- and off-CF maskers have been used in a forward-masking paradigm. Using high-level off-CF maskers may thereby lead to an overestimation of compression by as much as a factor of 2 [91]. Furthermore, physiological recordings in non-human mammals demonstrated that the amount of forward masking in the AN is not large enough to account for the behavioural forward masking, whereas physiological masking at the level of the inferior colliculus seems to be reflecting behavioural patterns [92]. Thus, behaviourally estimated BM I/O functions derived from forward masking paradigms may reflect mechanisms beyond cochlear processing. A modelling analysis as the one provided in the current study for evaluating the potential peripheral neural generators contributing to the EFRs might be useful also for exploring the contributing factors underlying level-growth functions obtained with, e.g., DPOAEs and psychoacoustical masking measures that have been used to estimate cochlear compression. For instance, cochlear transmission-line models [e.g., 93, 94] could be used to validate methods based on DPOAEs (or other oto-acoustic emissions), and an integration of the current AN model [44] with signal detection theory methods [e.g., 95, 96] could be applied to validate behavioural estimates of compression.

## Conclusion

The recorded EFR magnitude-level functions showed a compressive growth with slopes comparable to compression estimates using direct physiological recording in non-human mammals, and group-averaged results from psychoacoustical methods as well as DPOAE magnitude-level functions. Moreover, in the case of a mild threshold elevation, the estimated slopes were higher than in the case of normal hearing, also consistent with the interpretation of a less compressive response due to a reduction of cochlear gain. However, a test-retest analysis revealed that, while the individual EFR recordings showed good repeatability, the repeatability of the slope estimated from EFR magnitude-level functions was poor due to error propagation.

A computer model of the AN was used to simulate EFRs_AN_ magnitude-level functions. The model was able to correctly account for the saturation of the EFRs above 60 dB SPL observed at the carrier frequencies of 0.5, 1 and 2 kHz, and it was able to account for the single-slope monotonic growth at the highest carrier frequency of 4 kHz. A model analysis revealed that the compression values estimated from the simulated EFR_AN_ magnitude-level functions, obtained by summing up contributions from responding neurons across CF, was similar to the compression values estimated from the growth of the BM output, obtained at the on-CF cochlear segment. However, the model suggested that EFR_AN_ magnitudes at medium-to-high stimulus levels contained significant off-CF contributions which were the underlying mechanism leading to the compressive growth of the EFR_AN_ magnitude-level functions. Indeed, the on-CF contribution of simulated EFR_AN_ magnitude-level functions decreased with increasing level, which is at odds with the increasing compressive growth in the BM. In conclusion, despite the numerical match between the compressive growth of EFR magnitude-level functions and other estimates of cochlear compression using other methods, the model suggested that the compressive growth of the EFR magnitude-level functions may result from off-CF contributions, and not from the local on-CF compressive growth in the BM. This compromises the use of EFRs to estimate peripheral compression, together with the poor repeatability of the estimated slopes.

## Data Availability

The data reported in this article are publicly available in a Zenodo repository.

https://doi.org/10.5281/zenodo.844833

## Acknowledgements

The authors wish to thank to James M. Harte from Interacoustics Research Unit (IRU) for his valuable comments and discussions. We are also thankful to Ian C. Bruce for facilitating us with a version of the AN model that outputs the response of the BM module. This work was supported by the Oticon Centre of Excellence for Hearing and Speech Sciences (CHeSS) and the Novo Nordisk Foundation grant NNF17OC0027872 at the Technical University of Denmark (DTU).

## Author contributions statement

T.D. and B.E. conceived the experiments, G.E.L, T.D. and B.E. conceived the modelling analysis, G.E.L. conducted the experiments and modelling analysis, G.E.L analysed the experimental and modelling results. G.E.L prepared the figures. All authors reviewed the manuscript.

## Additional information

### Accession codes

(non applicable);

## Supplementary Information

**Supplementary Figure 1.**
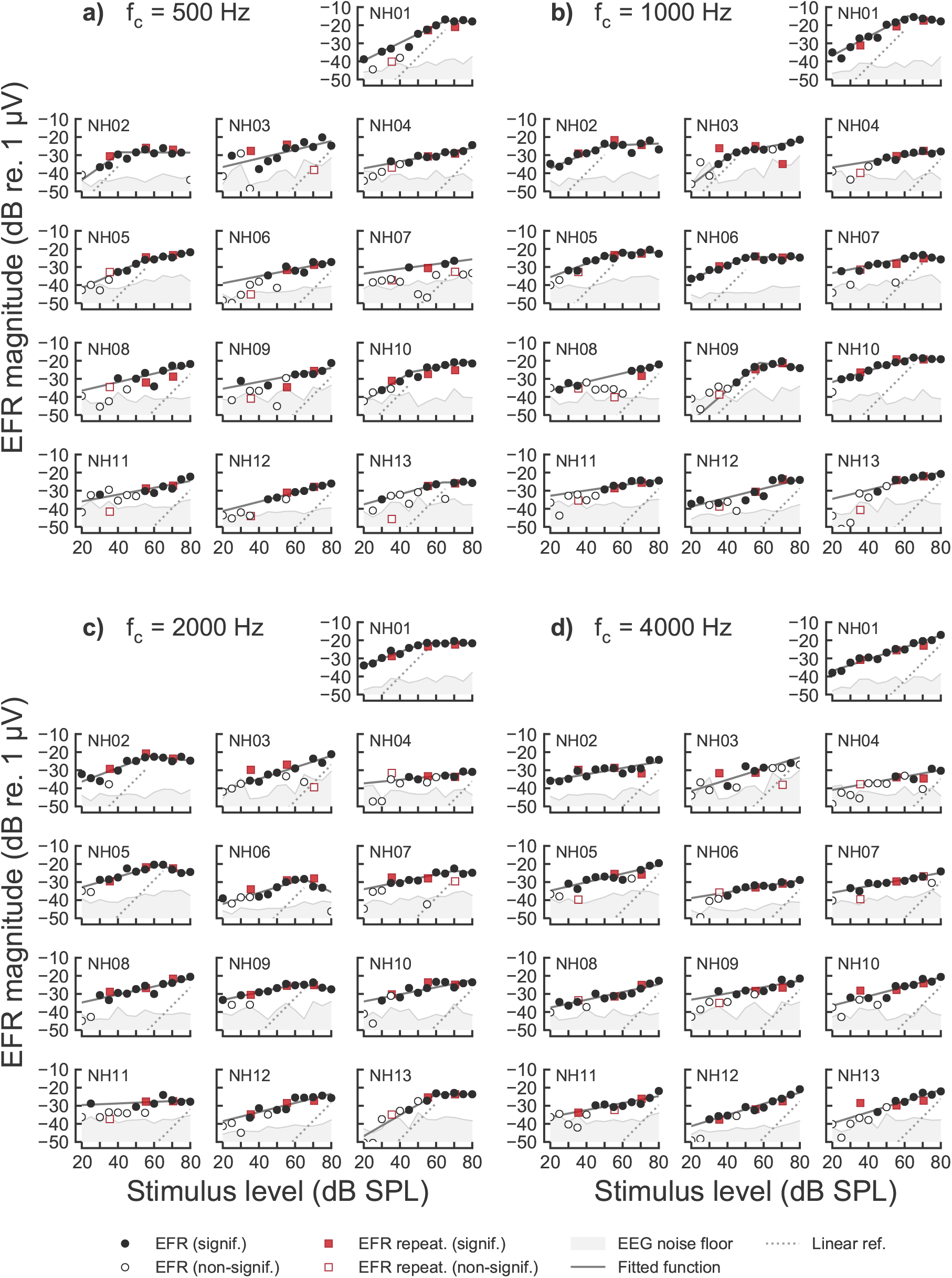
EFR magnitude-level functions recorded in all NH listeners for the carrier frequencies of a) 500 Hz, b) 1000 Hz, c) 2000 Hz and d) 4000 Hz. EFR magnitudes are represented as filled symbols in the case of a statistically significant response (positive F-test), and as open symbols in the case of statistically non-significant (negative F-test) responses. Circles indicate the EFR magnitudes recorded in the first recording session and red squares represent EFR magnitudes recorded in the second recording session. EEG background noise estimates are shown as the grey shaded area. The best fitted curves are represented by the solid dark-grey line. Linear reference with slope of 1 dB/dB is indicated by the dotted line.

**Supplementary Figure 2.**
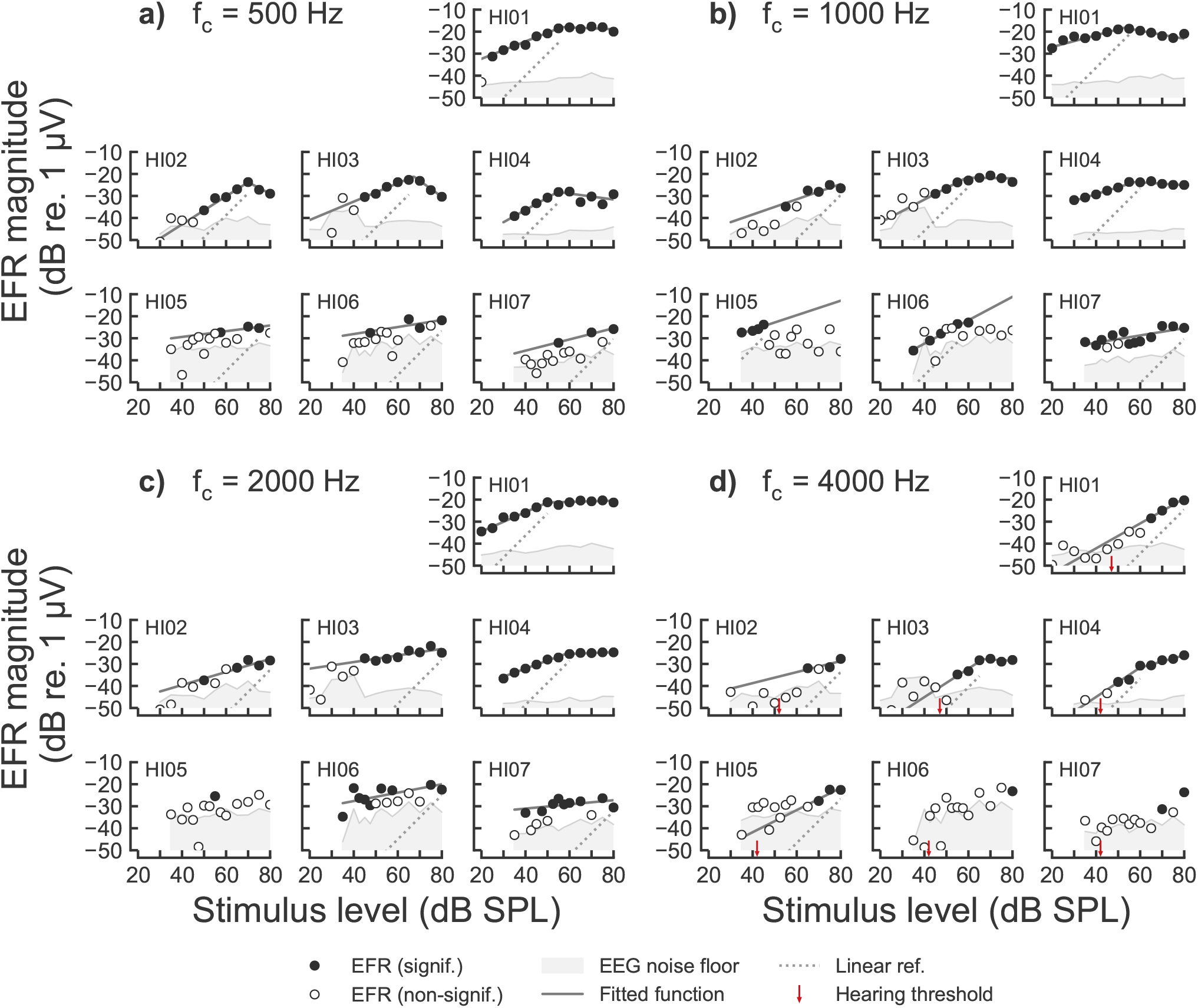
EFR magnitude-level function recorded in all HI listeners for the carrier frequencies of a) 500 Hz, b) 1000 Hz, c) 2000 Hz and d) 4000 Hz. Same representation as in Fig. 1 but not including the repeatability measurements. The small red arrow in panel d) indicates the behavioural hearing threshold of the listener at 4000 Hz in dB SPL.

**Supplementary Figure 3.**
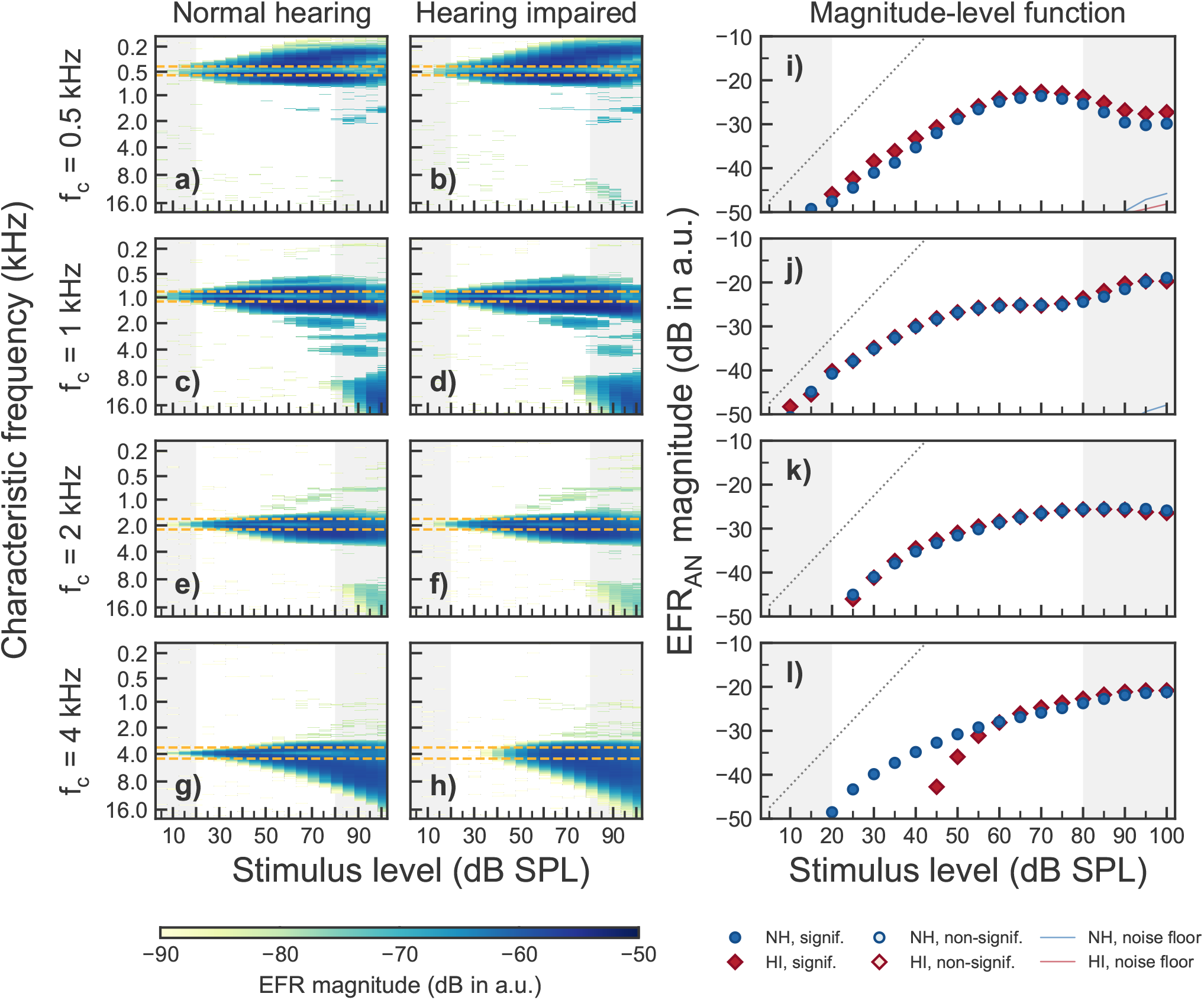
Simulated EFR at the level of the AN (EFR_AN_) obtained with four simultaneously presented SAM tones as the ones used in the experiments for NH and HI assuming only OHC dysfunction. Panels a-h show EFR_AN_ magnitudes as a function of stimulus levels for each of the four carrier frequencies (0.5, 1, 2 and 4 kHz). The colour gradient indicate significant EFR_AN_ magnitudes. The horizontal orange dashed lines indicate the on-frequency range as defined in the Results section in the article text. The vertical grey shaded areas show input level ranges outside the level ranges considered in the experiments. The two leftmost columns show simulated EFR for the mean audiogram of the NH (left) and HI listeners (middle), respectively. Panels i-j in the rightmost column show the corresponding EFR_AN_ magnitude-level functions obtained by summing up all AN activity across CF. Blue circles show NH simulations and red diamonds represent HI simulations. The thin solid lines represent noise floor estimates. Only OHC dysfunction was assumed to adjust the AN model parameters to account for the HI mean audiogram values.

**Supplementary Figure 4.**
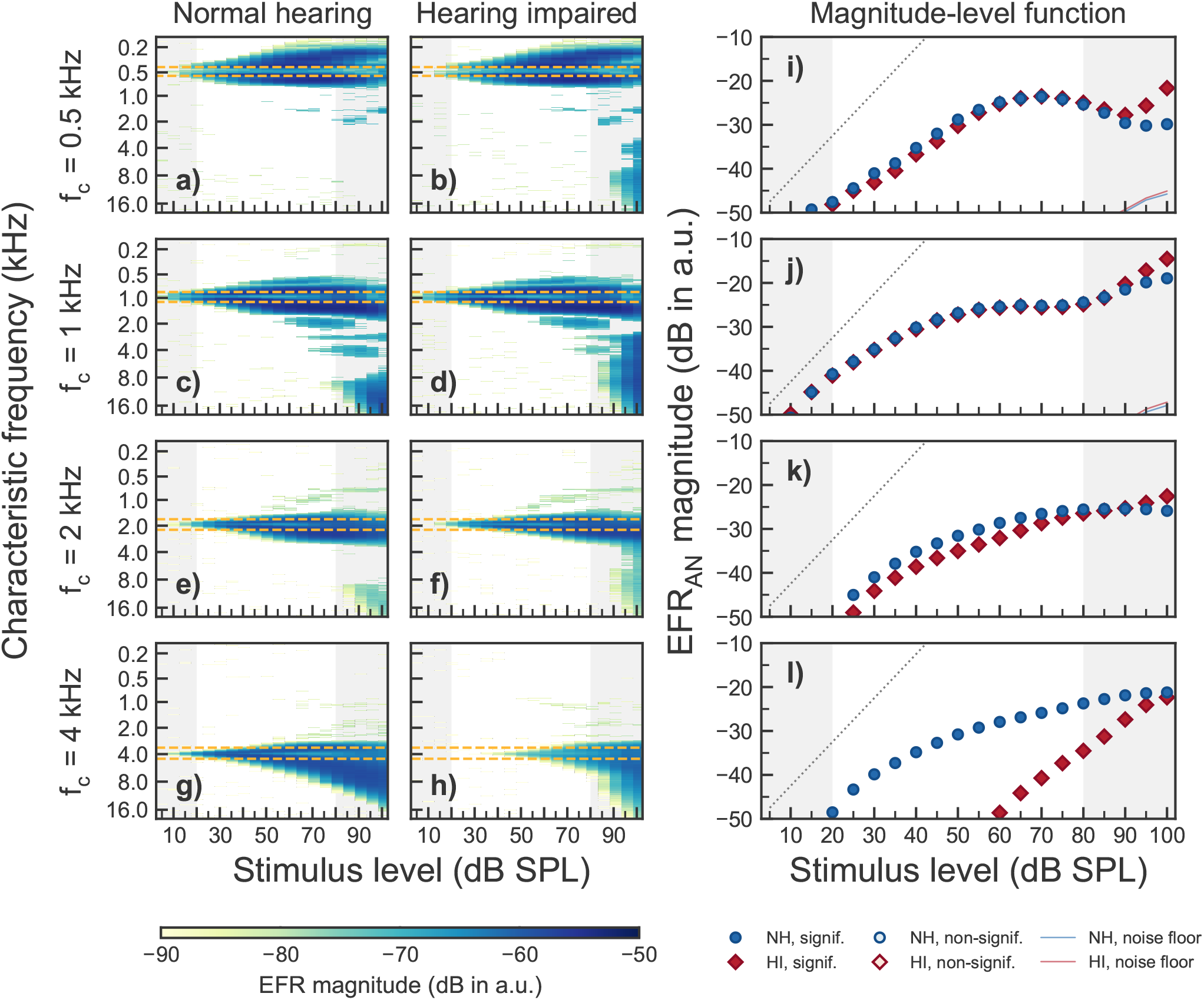
Simulated EFR at the level of the AN (EFR_AN_) obtained with four simultaneously presented SAM tones as the ones used in the experiments for NH and HI assuming only IHC dysfunction. Same representation as in Fig. 3 but assuming only IHC dysfunction to adjust the AN model parameters to account for the HI mean audiogram values.

**Supplementary Table 1.** Fitted parameters to the EFR magnitude-level functions for all NH listeners at all frequencies. The values in the brackets indicate the lower and upper 95% CI.

| Listener | Frequency |  |  |  |
| --- | --- | --- | --- | --- |
|  | 500 Hz | 1000 Hz | 2000 Hz | 4000 Hz |
| NH01 | $s_1 = 0.49$ (0.40, 0.57) | $s_1 = 0.52$ (0.40, 0.64) | $s_1 = 0.37$ (0.28, 0.46) | $s = 0.33$ (0.29, 0.37) |
| | $s_2 = -0.02$ (-0.47, 0.42) | $s_2 = -0.16$ (-0.57, 0.25) | $s_2 = -0.01$ (-0.13, 0.11) | $b = -43.5$ (-45.7, -41.3) |
| | $b_x = 65.0$ (54.1, 75.9) | $b_x = 61.8$ (53.4, 70.3) | $b_x = 53.8$ (46.6, 60.8) | |
| | $b_y = -18.6$ (-23.2, -14.1) | $b_y = -16.0$ (-19.8, -12.1) | $b_y = -22.4$ (-24.2, -20.6) | |
| NH02 | $s_1 = 0.71$ (-0.04, 1.46) | $s_1 = 0.42$ (0.17, 0.67) | $s_1 = 0.38$ (0.22, 0.54) | $s = 0.17$ (0.11, 0.23) |
| | $s_2 = -0.01$ (-0.21, 0.20) | $s_2 = 0.04$ (-0.16, 0.24) | $s_2 = -0.11$ (-0.43, 0.22) | $b = -39.2$ (-42.4, -36.0) |
| | $b_x = 41.7$ (31.6, 51.9) | $b_x = 50.0$ (33.9, 66.1) | $b_x = 57.0$ (44.6, 69.3) | |
| | $b_y = -28.4$ (-32.6, -24.3) | $b_y = -24.8$ (-28.8, -20.8) | $b_y = -22.2$ (-26.1, -18.4) | |
| NH03 | $s = 0.24$ (0.08, 0.40) | $s_1 = 0.77$ (0.12, 1.42) | $s = 0.32$ (0.24, 0.40) | $s = 0.31$ (-0.10, 0.72) |
| | $b = -41.3$ (-51.0, -31.7) | $s_2 = 0.18$ (0.11, 0.25) | $b = -47.6$ (-52.2, -42.9) | $b = -47.7$ (-71.3, -24.1) |
| | | $b_x = 43.0$ (35.8, 50.1) | | |
| | | $b_y = -28.9$ (-31.3, -26.4) | | |
| NH04 | $s = 0.19$ (0.12, 0.26) | $s = 0.16$ (0.07, 0.25) | $s = 0.10$ (0.00, 0.20) | $s = 0.18$ (-0.30, 0.67) |
| | $b = -41.1$ (-45.3, -36.8) | $b = -39.9$ (-45.7, -34.2) | $b = -39.3$ (-45.7, -32.8) | $b = -44.6$ (-78.1, -11.0) |
| NH05 | $s_1 = 0.45$ (0.11, 0.78) | $s_1 = 0.37$ (0.19, 0.56) | $s_1 = 0.29$ (0.15, 0.42) | $s = 0.24$ (0.15, 0.32) |
| | $s_2 = 0.17$ (0.05, 0.28) | $s_2 = 0.01$ (-0.23, 0.25) | $s_2 = -0.28$ (-0.60, 0.03) | $b = -39.5$ (-44.3, -34.7) |
| | $b_x = 55.0$ (40.7, 69.3) | $b_x = 56.0$ (42.7, 69.3) | $b_x = 53.8$ (46.6, 60.8) | |
| | $b_y = -27.0$ (-30.5, -23.6) | $b_y = -23.2$ (-27.1, -19.3) | $b_y = -21.6$ (-23.9, -19.3) | |
| NH06 | $s = 0.20$ (0.03, 0.37) | $s_1 = 0.38$ (0.30, 0.47) | $s_1 = 0.31$ (0.13, 0.49) | $s = 0.16$ (0.09, 0.24) |
| | $b = -42.9$ (-54.0, -31.9) | $s_2 = -0.02$ (-0.13, 0.09) | $s_2 = -0.48$ (-1.31, 0.35) | $b = -42.1$ (-46.8, -37.4) |
| | | $b_x = 51.6$ (45.2, 58.0) | $b_x = 64.9$ (56.1, 73.7) | |
| | | $b_y = -25.4$ (-27.2, -23.5) | $b_y = -29.3$ (-32.7, -25.8) | |
| NH07 | $s = 0.13$ (-0.39, 0.66) | $s_1 = 0.21$ (0.16, 0.25) | $s_1 = 0.20$ (0.09, 0.32) | $s = 0.18$ (0.13, 0.24) |
| | $b = -36.3$ (-68.3, -4.3) | $s_2 = -0.24$ (-0.47, -0.02) | $s_2 = -0.11$ (-0.76, 0.56) | $b = -39.6$ (-42.5, -36.6) |
| | | $b_x = 69.4$ (65.0, 73.7) | $b_x = 70.0$ (49.7, 90.3) | |
| | | $b_y = -23.2$ (-24.2, -22.2) | $b_y = -23.8$ (-26.9, -20.6) | |
| NH08 | $s = 0.24$ (0.01, 0.46) | $s = 0.24$ (0.19, 0.28) | $s = 0.22$ (0.14, 0.30) | $s = 0.21$ (0.15, 0.28) |
| | $b = -41.3$ (-55.3, -27.2) | $b = -41.1$ (-43.6, -38.6) | $b = -39.2$ (-43.7, -34.7) | $b = -41.8$ (-45.6, -38.0) |
| NH09 | $s = 0.19$ (0.06, 0.32) | $s_1 = 0.86$ (0.17, 1.55) | $s_1 = 0.21$ (0.14, 0.28) | $s = 0.17$ (0.06, 0.28) |
| | $b = -39.3$ (-47.7, -30.9) | $s_2 = -0.13$ (-0.44, 0.18) | $s_2 = -0.17$ (-0.39, 0.03) | $b = -36.7$ (-43.5, -29.9) |
| | | $b_x = 57.7$ (50.2, 65.2) | $b_x = 62.5$ (55.0, 70.0) | |
| | | $b_y = -22.1$ (-26.0, -18.2) | $b_y = -25.4$ (-26.9, -23.8) | |
| NH10 | $s_1 = 0.61$ (0.22, 1.01) | $s_1 = 0.31$ (0.26, 0.37) | $s = 0.20$ (0.10, 0.29) | $s = 0.26$ (0.16, 0.36) |
| | $s_2 = 0.18$ (0.02, 0.34) | $s_2 = -0.08$ (-0.25, 0.09) | $b = -38.1$ (-43.7, -32.5) | $b = -41.7$ (-47.5, -35.9) |
| | $b_x = 46.3$ (34.2, 58.5) | $b_x = 63.7$ (58.0, 69.4) | | |
| | $b_y = -26.2$ (-31.3, -21.1) | $b_y = -18.1$ (-29.6, -16.7) | | |
| NH11 | $s = 0.19$ (0.04, 0.34) | $s = 0.15$ (0.04, 0.26) | $s = 0.04$ (-0.07, 0.16) | $s = 0.19$ (0.05, 0.34) |
| | $b = -39.7$ (-49.2, -30.2) | $b = -35.7$ (-42.9, -28.6) | $b = -30.5$ (-37.8, -23.2) | $b = -40.0$ (-49.2, -30.9) |
| NH12 | $s = 0.26$ (0.23, 0.28) | $s = 0.25$ (0.14, 0.36) | $s = 0.25$ (0.14, 0.37) | $s = 0.31$ (0.25, 0.37) |
| | $b = -46.4$ (-48.0, -44.8) | $b = -43.8$ (-49.9, -37.7) | $b = -43.6$ (-50.7, -36.5) | $b = -47.5$ (-51.1, -43.8) |
| NH13 | $s_1 = 0.28$ (0.20, 0.35) | $s = 0.24$ (0.11, 0.38) | $s_1 = 0.58$ (0.38, 0.78) | $s = 0.31$ (0.19, 0.43) |
| | $s_2 = 0.00$ (-0.25, 0.24) | $b = -39.4$ (-48.2, -30.7) | $s_2 = 0.00$ (-0.20, 0.20) | $b = -45.8$ (-53.8, -37.9) |
| | $b_x = 63.6$ (51.9, 75.4) | | $b_x = 60.0$ (54.4, 65.6) | |
| | $b_y = -26.4$ (-29.3, -23.5) | | $b_y = -24.6$ (-27.3, -21.9) | |

**Supplementary Table 2.** Fitted parameters to the EFR magnitude-level functions for all HI listeners at all frequencies, including lower and upper 95% CI in brackets.

| Listener | Frequency |  |  |  |
| --- | --- | --- | --- | --- |
|  | 500 Hz | 1000 Hz | 2000 Hz | 4000 Hz |
| HI01 | $s_1 = 0.40$ (0.32, 0.47) | $s_1 = 0.27$ (0.16, 0.37) | $s_1 = 0.44$ (0.34, 0.54) | $s = 0.57$ (0.18, 0.95) |
| | $s_2 = -0.04$ (-0.17, 0.09) | $s_2 = -0.18$ (-0.32, -0.05) | $s_2 = 0.03$ (-0.05, 0.11) | $b = -64.8$ (-93.0, -36.5) |
| | $b_x = 56.5$ (50.9, 62.1) | $b_x = 52.7$ (45.7, 59.6) | $b_x = 50.0$ (44.2, 55.8) | |
| | $b_y = -17.9$ (-19.7, -16.1) | $b_y = -18.2$ (-19.9, -16.6) | $b_y = -21.3$ (-22.8, -19.8) | |
| HI02 | $s_1 = 0.64$ (0.35, 0.93) | $s = 0.35$ (0.00, 0.69) | $s = 0.30$ (0.04, 0.55) | $s = 0.25$ (-1.96, 2.46) |
| | $s_2 = -0.52$ (-1.83, 0.79) | $b = -52.3$ (-76.3, -28.3) | $b = -51.4$ (-68.7, -34.1) | $b = -48.8$ (-211.5, 114.0) |
| | $b_x = 70.0$ (60.6, 79.4) | | | |
| | $b_y = -24.8$ (-30.7, -18.9) | | | |
| HI03 | $s_1 = 0.42$ (0.31, 0.52) | $s_1 = 0.52$ (0.04, 0.99) | $s = 0.15$ (0.04, 0.26) | $s = 0.27$ (0.01, 0.52) |
| | $s_2 = -0.72$ (-0.96, -0.48) | $s_2 = -0.02$ (-0.23, 0.20) | $b = -35.2$ (-42.1, -28.2) | $b = -48.2$ (-65.7, -30.6) |
| | $b_x = 67.3$ (65.0, 69.6) | $b_x = 59.2$ (48.9, 69.5) | | |
| | $b_y = -22.2$ (-23.4, -20.9) | $b_y = -22.7$ (-25.3, -20.0) | | |
| HI04 | $s_1 = 0.56$ (0.11, 1.01) | $s_1 = 0.31$ (0.26, 0.37) | $s_1 = 0.41$ (0.36, 0.47) | $s = 0.41$ (0.26, 0.57) |
| | $s_2 = -0.14$ (-0.38, 0.10) | $s_2 = -0.07$ (-0.14, 0.01) | $s_2 = 0.04$ (-0.03, 0.12) | $b = -58.1$ (-68.3, -47.9) |
| | $b_x = 54.7$ (44.5, 64.8) | $b_x = 57.8$ (54.0, 61.5) | $b_x = 55.6$ (51.7, 59.6) | |
| | $b_y = -28.2$ (-31.4, -25.0) | $b_y = -23.7$ (-24.5, -22.8) | $b_y = -25.7$ (-27.0, -24.4) | |
| HI05 | $s = 0.13$ (-0.83, 1.09) | $s = 0.33$ (-0.11, 0.77) | | $s = 0.50$ (-3.32, 4.33) |
| | $b = -34.6$ (-99.8, 30.6) | $b = -39.3$ (-57.4, -21.3) | | $b = -61.9$ (-349.2, 225.4) |
| HI06 | $s = 0.16$ (-0.30, 0.61) | $s = 0.53$ (0.38, 0.68) | $s = 0.19$ (-0.02, 0.40) | |
| | $b = -34.4$ (-64.6, -4.2) | $b = -53.7$ (-61.0, -46.5) | $b = -35.3$ (-46.7, -23.9) | |
| HI07 | $s = 0.26$ (-0.31, 0.82) | $s = 0.17$ (0.06, 0.28) | $s = 0.10$ (-0.05, 0.24) | |
| | $b = -45.8$ (-85.0, -6.6) | $b = -38.7$ (-45.1, -32.2) | $b = -34.9$ (-43.4, -26.4) | |

